# The Impact of Internet Addiction on Academic Performance among Medical Students in Bangladesh: A Cross-Sectional Study and the Potential Role of Yoga

**DOI:** 10.1101/2023.06.28.23292017

**Authors:** Sumaiya Afrin, Nur-A-Safrina Rahman, Tahsin Tasneem Tabassum, Faisal Abdullah, Md. Istiakur Rahman, Sumona Haque Simu, Lakshya Kumar, Khutaija Noor, FNU Vishal, Vivek Podder

**Author notes:** Corresponding author: Sumaiya Afrin, Department of Health and Nutrition, Save the Children in Bangladesh, Dhaka, Bangladesh.

## Abstract

**Background:** Excessive internet use is a growing concern globally, and internet addiction negatively impacts academic performance. Limited research has been conducted on this topic among undergraduate medical students in Bangladesh.

**Purpose:** This study aimed to determine the prevalence of internet addiction and its impact on academic performance among medical students in Bangladesh, with a focus on the role of Yoga in reducing internet addiction.

**Methods:** A cross-sectional study was conducted among third-year medical students in four colleges in Dhaka, Bangladesh. Simple random sampling techniques were used to select participants, and data were collected using pre-tested questionnaires and a checklist for office records through face-to-face interviews. SPSS version 25 was used for data processing and analysis.

**Results:** Out of 312 students, 84% were addicted to the internet, and only 16% were normal. 64.4% had good results in the first professional examination, while 35.6% had poor results. 85.9% had good attendance, while 14.1% had poor attendance. The association between internet addiction level and first professional examination results and class attendance was highly significant.

**Conclusion:** This study highlights the high prevalence and negative impacts of internet addiction among medical students in Bangladesh. The study suggests that promoting awareness about healthy technology use, establishing a better balance between internet usage and academic study, and encouraging the practice of Yoga can help alleviate this problem. Further research and interventions are needed to tackle this emerging public health issue. Encouraging students to use the internet for academic purposes and providing education and resources can help promote healthy technology use. Healthcare professionals should also be aware of the risks and encourage seeking help if needed.

## Introduction

The WHO stated in 2014 that internet and smartphone usage have become integral to modern-day life and have globally increased in recent decades [1]. As of January 2021, around 4.6 billion people worldwide use the internet [2]. Internet addiction disorder was first proposed by Ivan Goldberg for pathological, compulsive internet usage [3]. Internet addiction disorder can be defined as “an individual’s inability to control his or her use of the internet, which eventually causes psychological, social, school, and/or work difficulties in a person’s life” [4]. With the increasing popularity and accessibility of the internet through various devices, internet addiction has become a concerning issue globally. Excessive use of the internet has been found to have negative consequences such as poor academic performance, relationship problems, and occupational difficulties [6, 7]. Internet addiction prevalence varies globally, with the UK and the US reporting 18.3% and 4.25% of college students being addicted, respectively [7]. In Qatar, India, and Taiwan, the addiction rates were 17.3%, 0.7%, and 17.9% among college students [7].

The internet has become a vital tool for knowledge acquisition and communication. The abundance of electronic books, encyclopedias, and dictionaries has made research more accessible and faster. Simulations, online slides, and presentations provide a more precise and elaborate view of topics. However, studies have shown that excessive use of social media can negatively impact academic performance [8]. Most scholars use social media for interaction, entertainment, and fun, which distracts them from academic work. Excessive social media use has been linked to poor academic performance, with scholars spending more time drooling and making musketeers than reading books [8, 9]. Employers have also suffered the negative effects of social media use on their employees [9]. Social media addiction can lead to poor academic performance and set back personal and professional responsibilities [9]. Social media platforms are more of a distraction than a tool for academic growth, with scholars spending more time on socializing conditioning than academic purposes. However, excessive internet use has been associated with academic problems and failure to complete assignments [10]. As of December 2018, 4.1 billion people worldwide were using the internet, with Asia having the highest number of users, led by China, India, and Bangladesh [11]. In Bangladesh, the number of internet users has exceeded 91.34 million people [12].

Research has indicated that while the internet has significant constructive benefits, excessive use and misuse can lead to addiction and pathological behavior [13]. Misuse of the internet, especially through excessive use, has resulted in a series of problems, including internet addiction disorder, which has attracted the attention of researchers worldwide [14, 15, 16]. Previous studies have reported that medical students use the internet mostly for non-academic purposes, such as entertainment and social networking, and that such use has negative effects on academic performance [17, 18, 19, 20, 21]. However, limited research has been conducted on internet addiction and academic performance among undergraduate MBBS medical students in Bangladesh. In Bangladesh, studies on the impact of internet use on academic performance have found mixed results. One study found a negative correlation between regular internet use and student results, while another identified several negative impacts on academic performance [22, 23]. However, over half of students’ surveyed reported improved academic performance due to internet use 24]. Another study found a positive correlation between study hours and internet use for academic purposes, while a negative correlation was found for non-study purposes [21]. Given the complexity and stress of academic course curricula, undergraduate medical students may be particularly susceptible to internet addiction [25], which could have negative consequences for their future career, lifestyle, and personality as doctors. Internet addiction has emerged as a major public health issue globally, and more and more students are becoming addicted to it, leading to detrimental effects on their health, studies, sleep, and family relationships [26].

Although a few studies have been conducted on internet addiction among medical students, results have varied widely due to differences in study design, assessment methods, and sampling from different sub-populations across the world. Therefore, this cross-sectional study aims to determine the prevalence of internet addiction and its impact on academic performance among undergraduate MBBS medical students in Bangladesh. The findings of this study could help raise awareness about internet addiction prevention and encourage students to take precautionary measures, as well as inform specific interventions by concerned authorities.

## Methods

### Study design

A cross-sectional study was conducted over a period of one year, from January 2021 to December 2021, in four medical colleges located in Dhaka, Bangladesh. The study population consisted of third-year medical students enrolled in the selected medical colleges, including Dhaka Medical College, Medical College for Women and Hospital, Tairunnessa Memorial Medical College, and International Medical College. The study places were selected based on convenience. Medical students from each college were selected by simple random sampling techniques, depending on their availability and willingness to participate.

### Data Collection

Data collection was carried out using pre-tested semi-structured questionnaires and a checklist for collecting office records.

### Data Collection Technique

Data were collected from respondents through face-to-face interviews after taking informed written consent.

### Target Population

Inclusion criteria must be:

1. An MBBS medical student, irrespective of gender,
2. Present in the college on the days of data collection,
3. Willing to participate by providing written informed consent.

Exclusion criteria must be:

1. Absent in the college on the days of data collection,
2. Unwilling to participate by providing written informed consent.

### Sample Size Estimation

The sample size was estimated using the following formula: n = z²pq/d²

Where,

n = the desired sample size

z = the value for 95% confidence level, usually set at 1.96

p = the proportion of internet addiction among medical students, which was reported to be 30.1% in a previous study (Zhang, 2018)

q = (1 - p) = 0.70

d = absolute precision, set at 5% (0.05)

Using these values, the desired sample size was calculated as: n = (1.96)² × 0.301 × 0.70 ÷ (0.05)² = 324

To minimize non-response after a 10% increase, the sample size was set at 356.

### Pre-Testing

Before data collection, a pre-testing of the questionnaire was conducted at Shaheed Monsur Ali Medical College. Data were collected from 20 respondents using a semi-structured questionnaire, and academic performance-related data were collected using a checklist. Based on the results of the pre-testing, no modifications were necessary, and the finalized questionnaire was used for data collection.

### Study Procedure

Data were collected with the help of students and administrative staff from the selected medical colleges. Permission to conduct the study was obtained from the medical college authorities with the submission of a request letter from Dhaka Medical College. Data were collected through face-to-face interviews using a pretested semi-structured questionnaire and checklist. Approximately 10-15 minutes were taken to collect data from each student. At the end of each interview, the collected questionnaires were checked for completeness and accuracy.

### Data Processing and Analysis

The collected data were checked, cleaned, edited, compiled, coded, and categorized according to the study objectives and variables to detect errors and maintain consistency, relevancy, and quality control. Data was entered into the computer for analysis, and the corrected data were analyzed using Statistical Package for Social Sciences (SPSS) version 25. Quantitative data were summarized by percentage, while qualitative data were summarized by mean and standard deviation.

### Ethical compliance

Prior to the commencement of this study, ethical approval of the research protocol was obtained from the Institutional Review Board (IRB) of Dhaka Medical College (DMC). This approval ensured that the study was conducted in an ethical manner, in accordance with relevant guidelines and regulations.

### Privacy and informed consent

It was ensured that the rights and welfare of the participants were protected. The aim and objective of the study, along with its procedure and benefits, were explained to the students in easily understandable local language and informed written consent was obtained. Each student was interviewed separately and the privacy and confidentiality of the respondents were maintained strictly. Any queries regarding questions and answers were clarified to the respondents as per their demand and desire. The respondents were informed about their full freedom to participate or refuse to participate in the study.

## Results

### Sociodemographic characteristics of respondents

The study used a semi-structured questionnaire to collect data on internet addiction, while academic data were collected using a checklist. A total of 312 students participated in the study. Table 1 summarizes the socio-demographic characteristics of the participants. The majority of the respondents were between 20-22 years old (95.5%), and the mean age was 21.05 ± 0.83 years. Most of the respondents were Muslim (83%), Bangladeshi (90.1%), and unmarried (94.2%). In terms of family structure, 78.0% of the respondents were living in a nuclear family, and the majority of the respondents (77.2%) were living in a hostel. In terms of monthly income, the majority of the respondents had a monthly income between BDT 20,000 to 40,000 (50.0%).

**Table 1:**
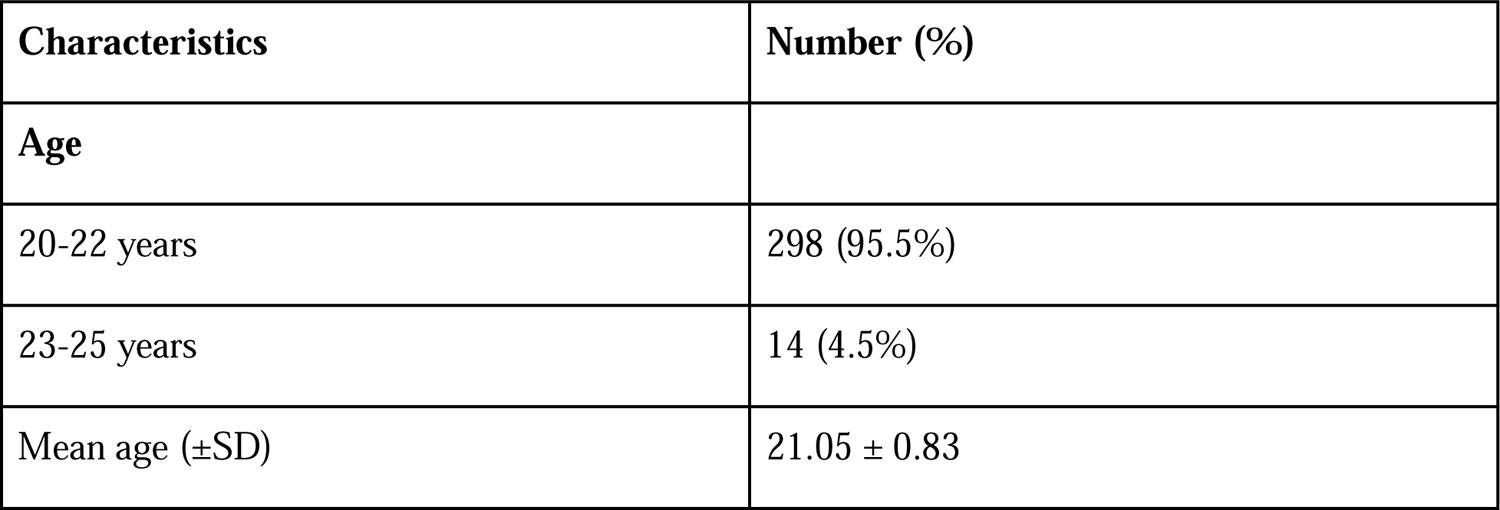
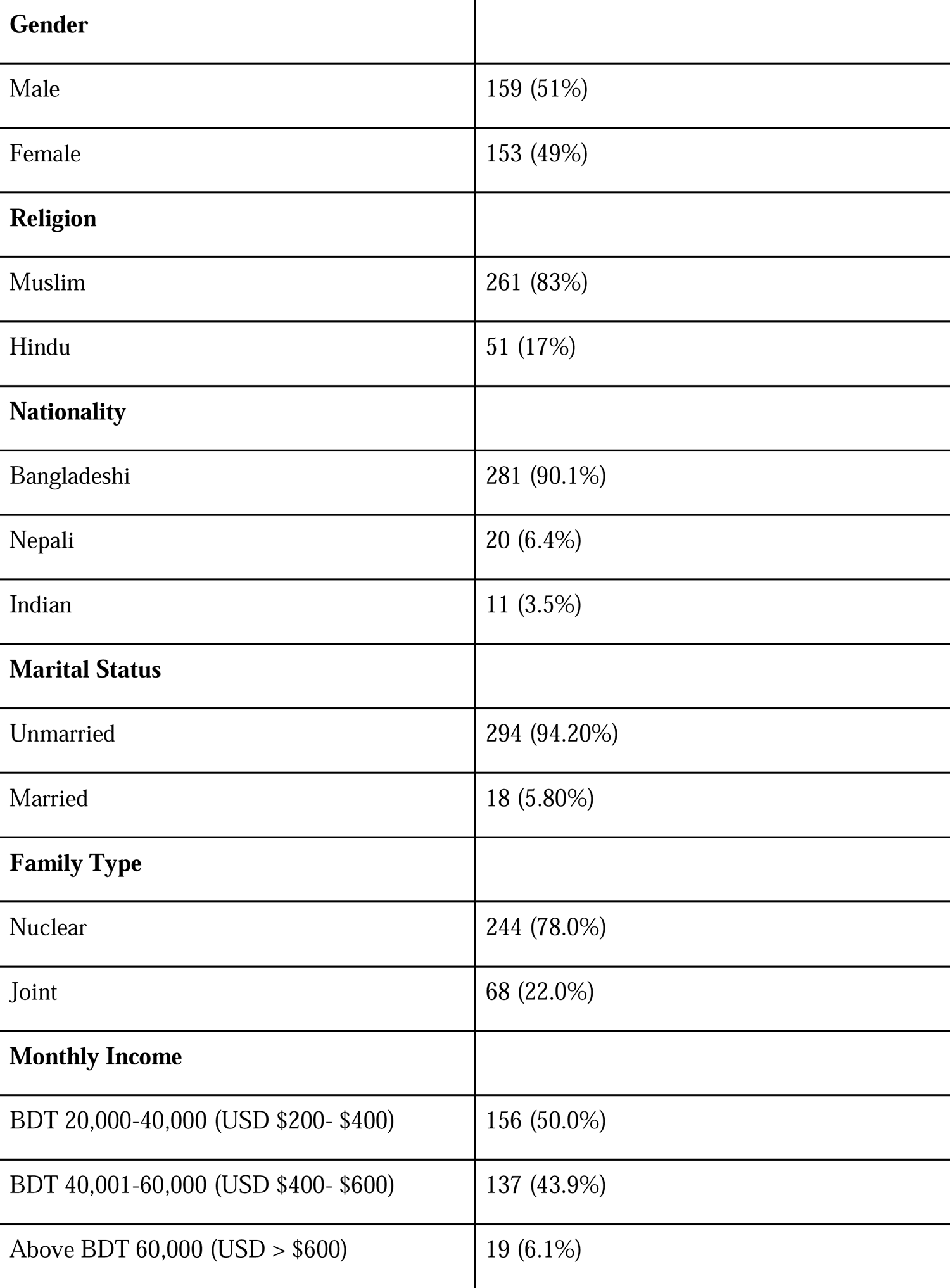
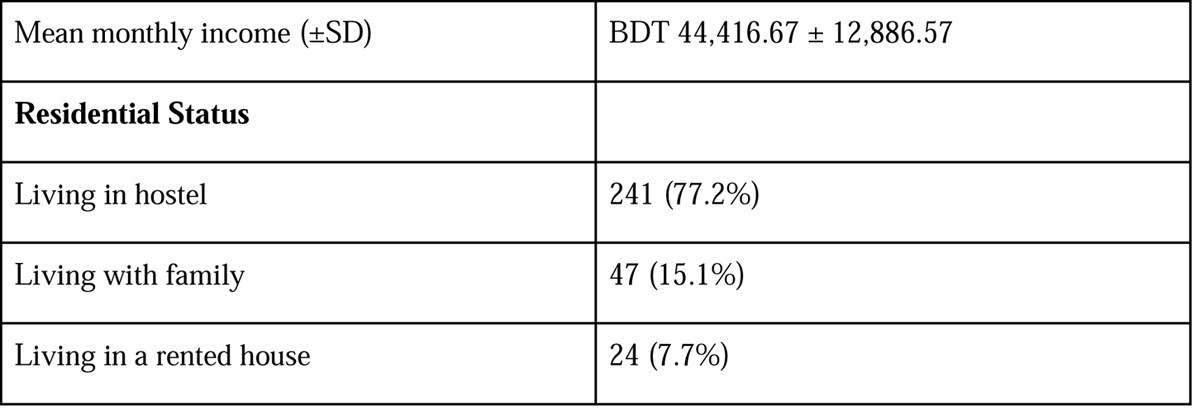
The socio-demographic characteristics of the participants

### Occupation of the respondents’ Parents

Table 2 shows the occupation of the respondents’ fathers, with service holder being the most common occupation (34.3%), followed by businessman (26.9%) and teacher (24.4%). The occupation of the respondents’ mothers is also shown in table 2 with housewife being the most common occupation (68.6%), followed by teacher (17.0%) and physician (6.1%).

**Table 2.** showing the occupation of the respondents’ parents

| **Occupation** | **Number (%)** |
| --- | --- |
| **Father** |  |
| Service holder | 107 (34.3%) |
| Businessman | 84 (26.9%) |
| Teacher | 76 (24.4%) |
| Physician | 27 (8.7%) |
| Farmer | 13 (4.2%) |
| Lawyer | 5 (1.6%) |
| **Mother** |  |
| Housewife | 214 (68.6%) |
| Teacher | 53 (17.0%) |
| Physician | 19 (6.1%) |
| Service holder | 19 (6.1%) |
| Nurse | 5 (1.6%) |

### Internet Addiction Status, Academic Performance, and Class Attendance

Table 3 shows that 84% of the students were addicted to the internet, with only 16% being normal. Among the addicted students, 36.9%, 35.6%, and 11.5% had mild, moderate, and severe addiction, respectively. In terms of academic performance, 64.4% of the respondents had good results in the first professional examination, while 35.6% had poor results. In terms of class attendance, 85.9% of the respondents had good attendance, while 14.1% had poor attendance.

**Table 3.** Internet Addiction, Academic Performance, and Class Attendance of Participants

| **Internet Addiction and Academic Performance** | **Number (%)** |
| --- | --- |
| **Internet Addiction Status** |  |
| Addicted | 262 (84%) |
| Normal | 50 (16%) |
| **Degree of Addiction (among addicted students)** |  |
| Mild | 115 (36.9%) |
| Moderate | 111 (35.6%) |
| Severe | 36 (11.5%) |
| **Academic Performance (First Professional Examination)** |  |
| Good (passed in all subject) | 201 (64.4%) |
| Poor results (failed in $\geq 1$ subjects) | 111 (35.6%) |
| **Class Attendance (Community and Forensic Medicine)** |  |
| Good attendance ($> 75\%$) | 268 (85.9%) |
| Poor attendance ($<75\%$) | 44 (14.1%) |

### Association between Internet addiction level and academic performance

Table 4 shows that among the students who had poor results in the first professional examination, 21.6%, 39.6%, and 34.3% were severely, moderately, and mildly addicted to the internet, respectively. Only 4.5% of these students were normal. Among the students who had good results in the first professional examination, 6.0%, 33.3%, and 38.3% were severely, moderately, and mildly addicted to the internet, respectively. About 22.4% of these students were normal. The association between the internet addiction level and first professional examination results was highly significant (P < 0.01).

**Table 4:** Association between Internet addiction level and academic performance and class attendance among students with good and poor attendance.

| Internet Addiction Level | Academic Performance |  | Class Attendance |  |
| --- | --- | --- | --- | --- |
|  | Good<br>(number [%]) | Poor<br>(number [%]) | Good<br>(number [%]) | Poor<br>(number [%]) |
| Normal | 45 (22.4%) | 5 (4.5%) | 48 (17.9%) | 5 (4.5%) |
| Mild | 77 (38.3%) | 36 (32.4%) | 103 (38.4%) | 12 (27.3%) |
| Moderate | 67 (33.3%) | 44 (39.6%) | 98 (36.6%) | 13 (29.5%) |
| Severe | 12 (6.0%) | 24 (21.6%) | 19 (7.1%) | 17 (38.6%) |
| **p value** | p< 0.01* |  |  |  |

\* p obtained from Chi square test

### Association between Internet addiction level and class attendance

Table 4 shows that among the students with poor class attendance, 38.6%, 29.5%, and 27.3% were severely, moderately, and mildly addicted to the internet, respectively. Only 4.5% of these students were normal. Among the students with good class attendance, 7.1%, 36.6%, and 38.4% were severely, moderately, and mildly addicted to the internet, respectively. About 17.9% of these students were normal. The association between the internet addiction level and class attendance was highly significant (P < 0.01).

## Discussion

The present study determined the prevalence of internet addiction and its relationship with academic performance among third-year MBBS medical students in Bangladesh. The majority of the participants had a mean age of 21 years, which was higher than in Nepal [27]. The sex distribution of the participants showed that males (51%) were slightly more represented than females (49%), which differed from a study in India where females (71.9%) were more represented [28]. The difference may be attributed to the availability of the male students during data collection. The majority of the participants were Muslim (83.0%). The majority of the participants were unmarried (94.2%), which is in line with a study in Nepal (90.7%) [27], but higher than in a Malaysian study (81.9%) [29]. This may be due to the fact that medical students in Bangladesh usually do not get married during their academic life.

In the current study, most of the participants belonged to a nuclear family (78%), which may indicate the rapid changing of family patterns in Bangladesh due to the expansion of education and employment opportunities, as well as westernization and modernization. The majority of the participants were living in a hostel (77.2%), which was higher than a study in India (65.96%) [30]. The prevalence of internet addiction among the study participants was 84.0%, which was in line with Pakistan (85%) [31] and Malaysia (81%) [29], but lower than in Egypt (87%) [32] and Nepal (92.8%) [27], and two studies in India (76.84% and 58.07%) [30, 33]. The prevalence of internet addiction among the study participants was 36.9% mild, 35.6% moderate, and 11.5% severe, while only 16% of students were not addicted to the internet. These findings differed from a study in Bangladesh [34] that reported 76.9% of the respondents had internet addiction, with 62.6% moderate, 13.1% mild, and 14.3% severe addiction. In an Indian study, the prevalence of internet addiction was 52.63% mild, 24.21% moderate, while 23.16% of students reported normal internet usage, and severe internet addiction was not reported among the participants [35]. The higher prevalence of internet addiction among the participants in this study may be due to increasing advances in technology and cheaper internet costs, as well as the COVID-19 pandemic leading to longer online periods and restricted outdoor activities.

The prevalence of internet addiction among the study participants was 84.0%, which was in line with Pakistan (85%) [31] and Malaysia (81%) [29], but lower than in Egypt (87%) [32] and Nepal (92.8%) [27], and higher than in Bangladesh (76.9%) [34] and two studies in India (76.84% and 58.07%) [30, 33]. The prevalence of internet addiction among the study participants was 36.9% mild, 35.6% moderate, and 11.5% severe, while only 16% of students were not addicted to the internet. These findings differed from a study in Bangladesh [34] that reported 76.9% of the respondents had internet addiction, with 62.6% moderate, 13.1% mild, and 14.3% severe addiction. In an Indian study, the prevalence of internet addiction was 52.63% mild, 24.21% moderate, while 23.16% of students reported normal internet usage, and severe internet addiction was not reported among the participants [35]. The higher prevalence of internet addiction among the participants in this study may be due to increasing advances in technology and cheaper internet costs, as well as the COVID-19 pandemic leading to longer online periods and restricted outdoor activities.

Furthermore, the current study revealed a significant negative association between internet addiction and academic performance. This finding is consistent with the other study conducted among medical students [36]. The negative association between internet addiction and academic performance could be attributed to several reasons. For instance, internet addiction may lead to time wastage, procrastination, distraction, and reduced attention span, which can adversely affect academic performance.

Yoga may benefit medical students in Bangladesh by reducing internet addiction and improving academic performance [37, 38, 39, 40, 41]. Yoga combines physical postures, breathing techniques, and meditation to enhance mental and physical health [37, 38, 39]. Studies have shown that yoga reduces stress, anxiety, and depression, which are common risk factors for internet addiction [37, 38]. Furthermore, yoga enhances self-control and self-regulation, decreasing the likelihood of internet addiction [38]. Practicing yoga also promotes mindfulness and attention, leading to improved focus on academic work [37, 38, 39, 40, 41]. Additionally, yoga increases cognitive function and memory retention by improving blood flow and oxygenation to the brain [37, 38, 39, 40, 41]. Including yoga in prevention and intervention programs can be cost-effective and accessible, benefiting medical students. Medical educators and policy-makers should consider incorporating yoga into the medical curriculum to promote its benefits.

The results of the present study have important implications for medical education and practice in Bangladesh. Given the high prevalence of internet addiction among medical students, it is crucial to develop and implement effective prevention and intervention programs such as yoga to reduce its negative consequences. Such programs may include raising awareness about the risks and symptoms of internet addiction, promoting healthy and balanced use of technology, providing counseling and psychological support services, and encouraging students to engage in physical activity, socialization, and extracurricular activities. Moreover, medical educators and policy-makers should recognize the impact of internet addiction on academic performance and consider incorporating relevant topics in the medical curriculum. This could include educating students about the impact of technology on health and well-being, providing strategies to manage internet addiction, and fostering critical thinking and digital literacy skills.

The present study has several limitations that should be considered when interpreting the results. Firstly, as a cross-sectional study, it cannot establish a causal-effect relationship between variables, and only provides the current prevalence of internet addiction among medical students in Bangladesh. Additionally, the study may have encountered selection bias as the sample was collected using a convenient sampling method. Further studies with larger and more representative samples and longitudinal designs are needed to confirm the findings and establish the directionality of the relationship between internet addiction and academic performance among medical students in Bangladesh.

## Conclusion

This study uncovers the concerning prevalence and negative effects of internet addiction among medical students in Bangladesh. The findings demonstrate that a significant portion of students struggle with internet addiction, resulting in poor academic performance and lower class attendance. To address this issue, it is recommended to build awareness among undergraduate medical students about the need to reduce internet addiction and promote healthy growth. In addition, incorporating yoga into interventions may offer a promising solution to reduce internet addiction among medical students in Bangladesh. Encouraging students to use the internet for academic purposes and establishing a better balance between internet usage and academic study is also important. Further research and interventions are necessary to tackle this emerging public health problem. Therefore, medical educators and policymakers should consider incorporating yoga practices into the curriculum and interventions for internet addiction among medical students. Additionally, healthcare professionals should be aware of the potential benefits of yoga and encourage patients to incorporate it into their daily routine.

## Data Availability

All data produced in the present study are available upon reasonable request to the authors.

## Acknowledgments

Not applicable.

## Authorship Contribution

Conceptualization: SA, NSR, TTT; Data curation: SA, NSR, TTT, FA; Formal analysis: SA; Investigation: SA, NSR, TTT; Methodology: SA, NSR, TTT, FA; Project administration: SA, NSR, TTT, FA, MIR; Software: SA, NSR; Supervision: SA, NSR; Validation: SA; Roles/Writing - original draft: SA, NSR, TTT, FA, MIR, VP; Writing - review & editing: SA, NSR, TTT, FA, MIR, VP, SHS, LK, KN, FV.

## ICMJE Statement

This article complies with the International Committee of Medical Journal Editors’ uniform requirements for the manuscript.

## Statement of Ethics

Prior to the commencement of this study, ethical approval was obtained from the Institutional Review Board (IRB) of Dhaka Medical College (DMC). This approval ensured that the study was conducted in an ethical manner, in accordance with relevant guidelines and regulations.

